# Association of *CYP11B2* T-344C polymorphism with hypertension and plasma aldosterone in a Ghanaian population

**DOI:** 10.64898/2026.07.11.26357813

**Authors:** Emmanuel Denu, Max Efui Annani-Akollor, Christian Obirikorang, Patrick Abankwah, Samuel Nkansah Darko

**Author notes:** Corresponding author (ED).

## Abstract

The Cytochrome P450 Family 11 Subfamily B Member 2 (*CYP11B2*) is the gene responsible for the synthesis of aldosterone synthase, the enzyme catalysing the terminal steps in the production of aldosterone. Polymorphism at the promoter region of this gene has been implicated to upregulate the synthesis of aldosterone synthase and the downstream production of aldosterone. A high aldosterone (hyperaldosteronism) is a known risk factor for hypertension. However, the association of the T-344C polymorphism with aldosterone production and the development of hypertension in the Ghanaian population has not been explored. Consequently, this study aims to unravel the relationship between T-344C (rs1799998) polymorphism, hypertension and serum aldosterone level. This study employed a case-control design enrolling 200 subjects of which 100 were hypertensive patients and 100 healthy controls in the Tamale Metropolis. Using a combination of genotyping, biochemical blood analysis and logistic modelling, we reveal that the frequency of the risk allele of rs1799998 (C) was higher amongst the patient group (0.565) than the control (0.315) group (p<0.0001). The adjusted (age and sex) logistic regression model revealed that the TC genotype [OR= 2.17 (1.08-4.38), p=0.0302] and the CC genotype [OR= 6.35 (2.60-15.54, p-value<0.0001] were significantly associated with hypertension. Under the genetic models, the recessive model (CC vs TC+TT) showed that the CC risk genotype was associated with hypertension [OR = 4.055 (1.838-8.949) p<0.001] after adjusting for age and sex. Furthermore, the CC risk genotype was associated with an elevated plasma aldosterone. In conclusion, the *CYP11B2* gene polymorphism (T-344C) was associated with high plasma aldosterone and hypertension.

## 1.0 Introduction

Cardiovascular diseases tend to have a high morbidity and mortality rate which is comparable to common diseases like tuberculosis, malaria and human immunodeficiency virus (HIV) in developing countries [1]. Hypertension lies in the centre of cardiovascular diseases: predisposing patients to stroke, heart attack, heart failure and arrhythmia [2]. With emerging problems of underdiagnoses, treatment and control of the disease [3,4], hypertension has become a public health burden across the world. Most research supports the notion that the disease is prevalent in developed countries [5,6], with comparable prevalence in urban areas in sub-Saharan countries. A recent cross-sectional study showed a 13 percent prevalence rate of hypertension among Ghanaians [7]. According to the World Health Organization [9], the normal blood pressure has a systolic reading of 90-120mmHg and a diastolic reading of 60-80mmHg. Studies suggests that lifestyle choices and socio-economic status influence hypertension development [4,8,9].

Nevertheless, the mediation of genetic factors in the onset and progression of hypertension cannot be underestimated and thus, studies on candidate genes that predisposes individuals have been documented. More than 20 genes have been discovered [10–12] to potentially mediate hypertension with over 100 single nucleotide polymorphisms. An important gene which has been implicated to influence blood pressure is the Cytochrome P450 Family 11 Subfamily B Member 2 (*CYP11B2*) gene. The gene codes for the aldosterone synthase enzyme responsible for catalysing the terminal steps in the production of aldosterone in a 3-step reaction pathway. Firstly, 11ß-hydroxylation of deoxycorticosterone (DOC) to corticosterone, 18-hydroxylation of corticosterone to 18-hydroxycorticosterone (18-OHB) and 18-oxidation of 18-hydroxycorticosterone to aldosterone [13]. This important mineralocorticoid (aldosterone) plays a vital role in maintaining fluid balance in the sodium-potassium pathway in the kidney. In the zona glomerulosa of the adrenal cortex, aldosterone binds to the mineralocorticoid receptor (MR) to cause the expression of the Epithelial Sodium Channel (ENaC) and activation of the sodium-potassium pump. This process causes the uptake of sodium and water into the distal renal and cortical collecting tubules, and excretion of potassium to the urine [14,15].

The *CYP11B2* gene is located at position 8q24.3 in the human chromosome [16]. At the promoter site of this gene, at position −344 is a single nucleotide polymorphism (SNP) which involves a base change from Thymine to Cytosine (T-344C) which has been associated to hypertension [17,18]. The site of this polymorphism represents the putative binding site for steroidogenic transcription factor-1 (SF-1) which induces transcription of the gene [19]. In the presence of the risk (C) allele, the affinity of the *CYP11B2* gene promoter to SF-1 increases and a corresponding increase in transcription [20– 22]. This produces more aldosterone synthase and a downstream increase in aldosterone than usual – activating water and sodium retention to increase blood pressure.

Genetic studies on SNPs are done in separate ethnicities or defined populations to identify associations in genetic markers [23] as the basis for establishing therapeutic targets. Americans with African ancestry have a high risk of hypertension with the −344T allele [24], which has no known reference to people living in Africa. Most of these studies were completed in Europe and Asia with conflicting results, suggesting a need to study the association of the rs1799998 polymorphism in diverse populations in Africa. In Ghana, there is a lack of literature on the association of this polymorphism on aldosterone production and the risk of hypertension development. Therefore, this study investigates the rs1799998 (T-344C) polymorphism in the *CYP11B2* gene and the risk it poses to hyperaldosteronism and the development of hypertension among Ghanaian subjects.

## 2.0 Materials and methods

### 2.1 Study participants and design

The study recruited 200 participants from the Tamale Metropolis in the Northern Region of Ghana. As the capital of this Metropolis, Tamale represents an urban city in Ghana, lying to the north between 9.16° and 9.34° latitude and, between 00.36°and 00.57 longitudes. The predominant occupations of the people in the Metropolis are trading and farming. Ethical clearance was obtained from the Ethics Committee of the University for Development Studies (UDS) Institutional Review Board with the reference number UDS/RB/009/22. Study participants were recruited on a voluntary basis from 1^st^ May 2022 to 31^st^ July 2022, and their consent were received by signing or thumbprinting on a consent form, after thoroughly explaining the research aims and procedure to them and agreeing to confidentiality and anonymity of data processing.

This case-control study involved 100 hypertensive patients and 100 normal controls. Participants from 25 to 70 years were recruited in the study and a case of hypertension was defined as a systolic and diastolic reading equal to or higher than 140/90 mmHg on three different occasions and these patients were already registered in the Tamale West and Tamale Central hospitals. Individuals with diabetes, hyperlipidaemia and congestive cardiac failure were excluded from the study. Control subjects were individuals with consistent blood pressure readings of 120/80 mmHg with no underlying conditions such as diabetes, hyperlipidaemia and congestive cardiac failure.

### 2.2 Anthropometric measurements

The weight of each participant was measured using the DETECTO 6855MHR (DETECTO, Webb City, MissouriUSA, 2020) series multifunctional scale, using recommended manufacturer protocol. Patients stood bare-footed, feet together, and standing in an upright position in 3 occasions and the average weight was recorded. The height rod was used to measure the height of participants, and the height and weight values were used to calculate the body mass index (BMI) of the participants. The BMI values were categorized into underweight (< 18.5 kg/m^2^), normal weight (18.5-24.9 kg/m^2^), overweight (24.9-kg/m^2^) and obese ((≥30.0 kg/m^2^), according to the world health organisation’s (WHO) definition.

### 2.3 Blood sampling

Participants were allowed to rest for 20 minutes upon arrival before their blood samples were collected. A total of 8 ml of venous blood was collected from each patient using a sterile syringe and subsequently divided into two 4 ml aliquots. One aliquot was stored in a sterile ethylene diamine tetra-acetic acid (EDTA) tube and the other part in a gel separator tube at −20°C. Content of the EDTA tube was used for DNA extraction for single nucleotide polymorphism analysis whereas the remaining sample in the gel separator tube was subjected to centrifugation (Centrifuge Z216MK, HEMLE Labortechnik GmbH) at 1500 RCF for 10 minutes and the separated serum was used for biochemical analysis.

### 2.4 DNA isolation and genetic analysis

The non-enzymatic salting-out method [25] was employed to isolate the genomic DNA. Briefly, the red and white blood cells were lysed using a low salt TKM1 buffer containing Triton X-100 and a high salt TKM buffer containing Triton X-100 respectively and the samples were incubated at 37°C for each digestion. The samples were centrifuged for 3 minutes at 5000 RCF in a tabletop centrifuge and the supernatant was discarded. A concentrated sodium chloride solution was used to precipitate the proteins out and a 100% isopropanol was used to further precipitate the genomic DNA. The precipitated DNA was dissolved in sterile water.

The rs1799998 polymorphism was investigated using restriction fragment length polymorphism (RFLP): Firstly, the promoter region of the *CYP11B2* gene was amplified with a forward primer: CAGGAGGAGACCCCATGTGAC and a reverse primer: CCTCCACCCTGTTCAGCCC through polymerase chain reaction (PCR). A PCR reaction volume of 25µL, containing the OneTaq® 2X Master Mix (New England Biolabs), 0.15µM of primer and 2ul of genomic DNA template was subjected to 35 cycles of reaction including an initial denaturation step at 94°C for 60 seconds, followed by the 35 cycles of 30-seconds denaturation at 94°C, 30 seconds annealing at 58°C, 45 seconds extension at 72°C and a final extension for 5 minutes at 72°C. The 538-base product was cleaved with the HaeIII restriction enzyme (New England Biolabs) for 13 minutes at 37°C, followed by enzyme inactivation for 20 minutes at 80°C. The digested products were subjected to a 2% agarose gel electrophoresis. The wild type (−344T) yielded a 274bp fragment. The presence of the polymorphism (−344C) creates an additional restriction site within the 273bp fragment, to produce 203 bp and 71 bp fragments. Thus, 274bp (T-allele) and 203bp (C-allele) fragments were detected.

### 2.5 Aldosterone measurement

Aldosterone level was quantified from serum samples using Enzyme-Linked Immunosorbent Assay (ELISA). The **DirectX**^**®**^ Aldosterone ELISA kit was used following manufacturer’s protocol. The aldosterone level was expressed as picogram per milliliter (pg/mL).

### 2.6 Statistical analysis

The demographic data of participants, biochemical data and genotyping results were entered into the Statistical Package for Social Sciences (SPSS v30) and GraphPad Prism (v8). Continuous variables were analysed using the student t-test and categorical data were analysed using Chi-square test. Where comparisons between more than two groups were performed, the analysis of variance (ANOVA) test was used. For the comparisons, a p-value < 0.05 was considered statistically significant. A logistic regression was fitted to determine the association of the polymorphism to hypertension; the SNPs were presented as predictor variables where odds ratio (OR) and 95% confidence intervals were reported. Since age and sex are risk factors for hypertension and these parameters were different amongst the study groups, models were adjusted for age and sex.

## 3.0 Results

### 3.1 Demographic and physiological data

The summary of demographic and physiological data of the study groups: cases (patient group) and controls Table 1 indicate that the patient group were older than the control group (51.76 vs 44.49, p=0.002) and the systolic blood pressure (mmHg), diastolic blood pressure (mmHg) and heart rate (bpm) were all higher in the patient group compared to the control group (p-value <0.05). There were more female participants in the study; making up 84% in the patient group, and 58% in the control group. There was no difference in the weight status between the study groups.

**Table 1.** Descriptive summary of demographic and physiological data.

| Variables | Patient Group | Controls | P-value |
| --- | --- | --- | --- |
|  | (N=100) | (N=100) |  |
| Age (years) | 51.76±16.45 | 44.49±16.34 | 0.0020 |
| BMI (Kg/m<sup>2</sup>) | 26.71±6.26 | 24.68±4.48 | 0.0092 |
| Sex |  |  | <0.0001 |
| Male | 16% | 42% |  |
| Female | 84% | 58% |  |
| Physiological Data |  |  |  |
| Systolic BP (mmHg) | 147.85±22.26 | 125.23±17.12 | <0.0001 |
| Diastolic BP (mmHg) | 89.59±11.18 | 78.48±9.57 | <0.0001 |
| Heart Rate (bpm) | 82.88±12.65 | 78.68±14.66 | 0.0312 |
| Weight Status |  |  | 0.5715 |
| Normal/Underweight | 47% | 51% |  |
| Overweight/Obese | 53% | 49% |  |

Summary of demographic and physiological data are presented as mean±std or proportions. Continuous data was analysed using the student t-test whereas the categorical data was analysed using Chi-square test. A p-value < 0.05 was considered significant

### 3.2 Genotypic and allelic frequencies of *CYP11B2* gene polymorphism in the study participants

The genotypic and allelic frequencies of the *CYP11B2* (T-344C) polymorphism in the study groups Table 2 revealed a higher minor allele frequency (C) in the patient group (0.565) than the control group (0.315) indicating the patient group has a higher frequency of the polymorphism and the difference was significant (p<0.0001) whereas the major allele frequency was significantly higher in the control group (0.685) compared to the patient group (0.315). The homozygote recessive genotype (CC) was predominant in the patient group (34%) than the controls (11%) whereas the homozygote dominant genotype (TT) was higher in the control group (48%) compared to the patient group (21%) and the Hardy-Weinberg equilibrium was observed in the control group (p=0.9485).

**Table 2.** Distribution of the *CYP11B2* gene polymorphism among the patient and control groups.

| SNP | Patient Group (n=100) |  |  | Control (n=100) |  |  |  |
| --- | --- | --- | --- | --- | --- | --- | --- |
|  | N | Allele frequency (N) |  | N | Allele frequency (N) |  | P-value |
|  |  | T | C |  | T | C |  |
|  |  | 0.435 (87) | 0.565 (113) |  | 0.685 (137) | 0.315 (63) | <0.0001 |
| TT | 21 |  |  | 48 |  |  | <0.0001 |
| TC | 45 |  |  | 41 |  |  |  |
| CC | 34 |  |  | 11 |  |  |  |
| **HWE-P** |  |  |  |  |  |  | 0.9485 |

The allelic and genotypic frequencies are presented as counts, and the Chi-square test was used to analyse the data with a p-value < 0.05 considered as significant.

### 3.3 Association between *CYP11B2* genetic polymorphism (rs1799998) and Hypertension

The risk of individuals with heterozygous TC and the homozygous recessive (CC) genotypes to develop hypertension was determined using a logistic regression where the TT dominant genotype was used as a reference. After adjusting for age and sex, the TC genotype [OR= 2.17 (1.08-4.38), p<0.05] and the CC genotype [OR= 6.35 (2.60-15.54), p-value<0.0001] were significantly associated with hypertension Table 3.

**Table 3.** Genotypic association of the *CYP11B2* polymorphism (T-344C) and hypertension.

| SNP | cOR (95% CI) | aOR (95% CI) | P-value |
| --- | --- | --- | --- |
| TT | Reference | Reference |  |
| TC | 2.51 (1.29-4.88) | 2.17 (1.08-4.38) | 0.0302 |
| CC | 7.07 (3.02-16.56) | 6.35 (2.60-15.54) | < 0.0001 |

Logistic regression using the TT dominant genotype as the reference to compare the TC and CC genotypes. The results are presented as odds ratio and confident intervals. Where cOR= Crude model and aOR= adjusted model for age and sex and a p-value <0.05 was considered significant.

A crude genetic model was fitted and since age and sex are risk factors to hypertension, a separate model adjusted for age and gender was fitted. Under the various genetic models, the recessive model (CC vs TC+TT) revealed that individuals with the homozygous recessive genotype stands a four-fold risk (cOR= 4.168) of hypertension Table 4 and the association was not weakened

**Table 4.** The genotypic model of *CYP11B2* polymorphism and hypertension.

| Model | Genotype | cOR* (95% CI) | p-value* | aOR**(95% CI) | p-value |
| --- | --- | --- | --- | --- | --- |
| Dominant | (TC+CC) vs TT | 3.473 (1.866-6.461) | <0.0001 | 3.042 (1.581-5.854) | 0.0009 |
| Recessive | CC vs (TC+TT) | 4.168 (1.967-8.830) | 0.0002 | 4.055 (1.838-8.949) | 0.0005 |
| Overdominant | TC vs (TT+CC) | 1.177 (0.672-2.062) | 0.5679 | 1.045 (0.576-1.898) | 0.8839 |

Inheritance models of the genotypes and the data are presented as odds ratio and confidence intervals. A p-value < 0.05 was considered significant.

after adjusting for age and sex (aOR= 4.055). In the dominant model, individuals with at least a single copy of the risk allele (C) had a chance of developing hypertension (cOR = 3.473 and aOR=3.042).

### 3.4 Relationship between the T-344C polymorphism and the physiological characteristics of the study groups

The distribution of clinical data across the various genotypes (TT, TC and CC) of the T-344C polymorphism Table 5 revealed no significant difference between the genotypes for BMI (p=0.9816) and heart rate (p=0.1202). However, The systolic blood pressure (p=0.0004) and the diastolic blood pressure (p=0.0002) were significantly higher in individuals with the homozygous recessive genotype (CC) than individuals with the homozygous dominant (TT) and heterozygous (TC) genotypes.

**Table 5.** The distribution of clinical data across the various genotypes (TT, TC and CC).

| Characteristics | TT | TC | CC | P-values |
| --- | --- | --- | --- | --- |
|  | (N=69) | (N=86) | (N=45) |  |
| BMI (kg/m<sup>2</sup>) | 25.73±6.88 | 25.74±4.80 | 25.55±4.52 | 0.9816 |
| Systolic BP (mmHg) | 130.25±22.32 | 135.94 ±20.63 | 147.32±24.12 | 0.0004 |
| Diastolic BP (mmHg) | 80.82±8.20 | 83.56±12.71 | 89.86±12.66 | 0.0002 |
| Heart Rate (bpm) | 78.59±14.77 | 80.84±13.06 | 84.03±13.36 | 0.1202 |

A one way ANOVA was used to compare the physiological characteristics amongst the genotypes. The results are presented as mean±std and a p-value < 0.05 was considered significant.

### 3.5 Aldosterone levels correlate with hypertension and the CC recessive genotype

A high aldosterone level is a clinical factor for hypertension, and the aldosterone synthase gene polymorphism (T-344C) has been implicated in the abnormally high production of aldosterone. Consequently, aldosterone level was measured amongst the study groups. The results indicate that there was a high aldosterone level in the patient group compared to the control group Fig 1A. Since the T-344C polymorphism has been implicated in the increase of aldosterone production, comparison of aldosterone levels among the genotypes were performed. The aldosterone levels in the heterozygous TC group increased (160.56±59.66) compared to the homozygous dominant (TT) group (103.421±23.83), whereas the homozygous recessive (CC) genotype showed a remarkably high aldosterone level (265.86±60.68) and the ANOVA test shows a significant difference between the genotypes (P<0.0001), suggesting that the −344C allele is associated with high aldosterone and hypertension Fig 1B.

**Fig 1.**
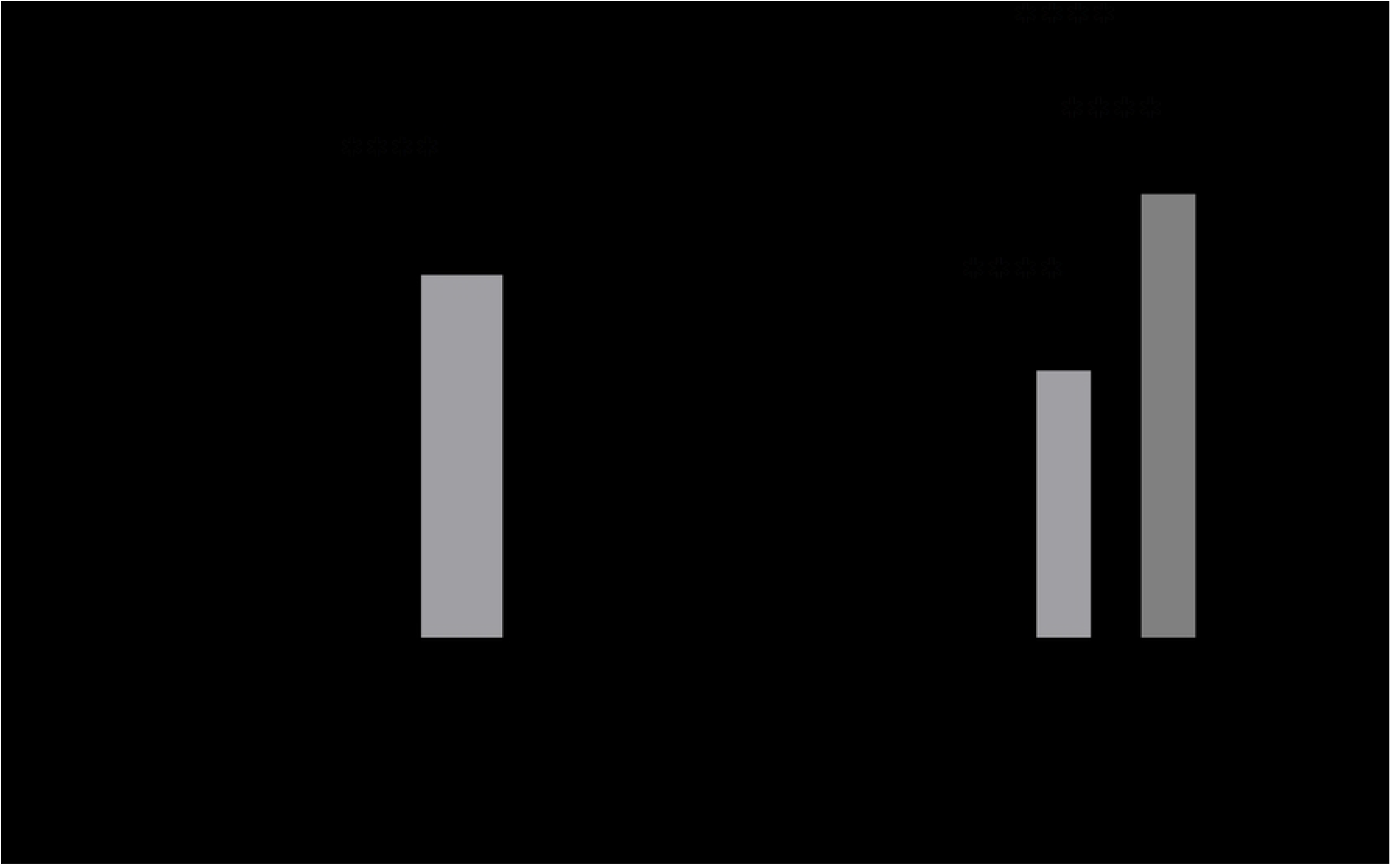
The CC genotype is associated with elevated aldosterone. A) Comparison of aldosterone level amongst patients and controls B) Aldosterone levels in the study group according to the genotypes. Statistical tests were performed using student t-test (A) and ANOVA (B) and data are presented as mean, and the error bars are represented as standard error of mean. **** means P < 0.0001.

## 4.0 Discussion

Polymorphisms in the *CYP11B2* gene have been studied in association with increasing transcriptional activity of the gene. The polymorphism and the associated hypertension and aldosteronism have been assessed in many populations with conflicting results [17,26,27]. Thus, this study assessed the rs1799998 polymorphism in the Ghanaian population for the first time to ascertain its influence on hypertension and aldosterone production.

The study revealed that the minor allele frequency of rs1799998 (C) was significantly different in the two groups studied (patient group and controls). The risk allele (C) was found in 31.5% of the control participants and 56.5% of the patient group, indicating a correlation of the T-344C base change to hypertension, which is confirmed by the genotypes, where the heterozygous (CC) genotype was found in 11% of the control group and 34% of the patient group. The heterozygous (TC) and the homozygous (TT) genotypes were present in 41% and 48% of the control group in comparison with 45% and 21% of the patient group, respectively. A previous study in an Anglo-Celtic population [18] revealed similarly high number of the risk (C) allele in the patient group (52%) than the control group (42%) and for the TC and TT genotypes, the outcome was like our findings: with 47% and 24% respectively in the patient group. Studies in other populations revealed similar results to this present study. A higher level of the risk allele (C) in the patient (case) group compared to the controls were reported in the Chinese population 41.1% vs 31.7% [27] and 34.2% vs 26.5% in a Japanese population [17]. However, studies in other populations have reported a higher level of the (C) allele in the disease-free control group, implicating the (T) allele as the risk allele in such populations. For example, in an Egyptian population, there is a higher level of the (C) allele amongst the controls (54.0%) compared to the patient group (34.5%) [26].

In the recessive genetic model, patients with the CC genotype have a 4-fold risk of developing hypertension compared with individuals with the TC and the TT genotypes, suggesting that the CC risk genotype is associated with hypertension in the study group. In the putative binding site of SF-1 transcription factor at the promoter region of the *CYP11B2* gene, a study has revealed that the T allele decreases the binding affinity of the gene to the SF-1 transcription factor by 4-fold and the mechanism of the gene regulation has been extensively discussed [19]. The increased binding affinity by the C allele upregulates the transcription rate of the gene to produce more aldosterone and a downstream increase in blood pressure.

A report from a European population [28] revealed an association between the CC genotype and an increased aldosterone level in hypertensive patients. Our present results confirm these previous studies that the CC genotype is associated with high aldosterone production. Nevertheless, there was no association between the genotypes of rs1799998 and plasma aldosterone in another study involving people of European descent [29], unravelling the dynamic differences in how the rs1799998 polymorphism affect aldosterone production and hypertension, requiring more work to be done in different populations to uncover the regional differences on the effect of this polymorphism on aldosterone production, and the onset and progression of hypertension.

## 5.0 Conclusion

The present study showed that the rs1799998 polymorphism is associated with hypertension and high aldosterone production. The CC genotype significantly increases the risk of developing hypertension and high aldosterone production in the study population.

## 6.0 Limitations

The study measured serum aldosterone levels which are critical in assessing the influence of the T-344C polymorphism. However, clinically, there is a combination of aldosterone and rennin levels into a ratio of aldosterone to rennin to screen for secondary hypertension. Thus, the study could have included the rennin levels to increase the robustness of the study.

## Data Availability

The minimal data is provided in the manuscript and the supporting files

## Acknowledgement

The authors acknowledge the Department of Molecular Medicine in the Kwame Nkrumah University of Science and Technology for providing the resources to complete this study.

## Supporting information

S1 Checklist. **Strobe checklist**.

S2 Data. **Minimal data**.

